# LinkR: an open source, low-code and collaborative data science platform for healthcare data analysis and visualization

**DOI:** 10.1101/2024.07.03.24309872

**Authors:** Boris Delange, Benjamin Popoff, Thibault Séité, Antoine Lamer, Adrien Parrot

## Abstract

**Background:** The development of Clinical Data Warehouses (CDWs) has greatly increased access to big data in medical research. However, the lack of standardization among different data models hampers interoperability and, consequently, the research potential of these vast data resources. Moreover, data manipulation and analysis require advanced programming skills, a skill set that healthcare professionals often lack.

**Methods:** To address these issues, we created an open source, low-code and collaborative data science platform for manipulating, visualizing and analyzing healthcare data using graphical tools alongside an advanced programming interface. The software is based on the OMOP common data model.

**Results:** LinkR enables users to generate studies using data imported from multiple sources. The software organizes the studies into two main sections: individual and population data sections. In the *individual data section*, user-friendly graphical tools allow users to customize data presentation, recreating the equivalent of a medical record, according to the needs of their study. The *population data section* is designed for conducting statistical analyses through both graphical and programming interfaces. The application also incorporates collaborative features, such as a messaging page and an integrated Git module. These features facilitate efficient collaboration and shared data analysis efforts across different research centers.

**Conclusion:** LinkR is a low-code data science platform that democratizes access, manipulation, and analysis of data from clinical data warehouses and facilitates collaborative work on healthcare data, using an open science approach.

## Background

The development of Electronic Health Records (EHRs) and Clinical Data Warehouses (CDWs) in recent years has enabled access to big data in the field of medical research (1). However, the lack of standardization across data models has been a significant barrier to interoperability and, consequently, to the reproducibility of research derived from these CDWs (2). To address this issue, Common Data Models (CDMs) such as the Observational Medical Outcomes Partnership (OMOP) by OHDSI (Observational Health Data Sciences and Informatics) and the Fast Healthcare Interoperability Resources (FHIR) have been developed (3–5). These initiatives are part of an open science approach, aiming the sharing of tools, methods and analyses conducted on health data (6).

Once the Extract-Transform-Load (ETL) process is completed, which implements data from EHRs into CDWs, analyses are typically conducted using tools such as RStudio and Jupyter Notebooks (7,8). While these softwares are powerful for data analysis, offer robust data visualization capabilities and facilitate easy communication of results, they require advanced programming skills, a competence usually absent in medical training. Healthcare professionals often lack the knowledge and experience to manipulate data and perform statistical analyses by themselves (9). To bridge this knowledge gap, some software solutions provide a “no-code” approach with graphical interfaces, such as SAS Enterprise Guide or IBM SPSS Statistics (10,11). However, these proprietary solutions provide limited functionalities for data visualization and analysis, and lack the flexibility for incorporating new features or customizations.

OHDSI’s ATLAS software, an open-source web-based tool, provides a functional platform for OMOP-format healthcare data visualization and analysis. It integrates with HADES, a set of open source R packages, enhancing its capabilities for comprehensive observational studies (12). Despite its strengths, ATLAS has limitations, including a restricted ability to add new functionalities, a lack of support for direct R or Python programming, and limited visualization options for individual patient records.

The objective of our study was to develop a modular, open source, low-code and collaborative data science platform. This platform is designed to facilitate healthcare data research, combining user-friendly graphical tools with an advanced programming interface for comprehensive visualization and analysis of healthcare data.

## Methods

### Specifications

Our aim was to develop an open source, low-code and collaborative data science platform for healthcare professionals, data scientists and healthcare students.

The software had to support data import from a variety of sources, including Excel, CSV and Parquet files, as well as relational databases. To ensure interoperability, it was imperative that the data processed within the application conformed to a standard format such as the OMOP CDM. The software had to provide both graphical tools for users with limited programming skills and a programming interface for more advanced data manipulations. The software had to enable any type of studies to be carried out, whether using classical statistics or machine learning methods. It had to be possible to add new functionalities to the application, such as graphical interfaces for creating figures or generating statistics from data. In line with open science principles, all work carried out on the application had to be shareable. The specifications are reported in *Table 1*.

**Table 1.** Software specifications.

| Feature | Description |
| --- | --- |
| Import data from various sources | Import Excel, CSV and Parquet files, import data from relational databases |
| Use a common data model | Process data using a common data model, such as OMOP or FHIR |
| Graphical user interface | User-friendly interface for data manipulation and visualization without coding |
| R programming interface | Ability to program and execute R scripts within the platform |
| Python programming interface | Ability to program and execute Python scripts within the platform |
| Patients chart review | Facilitate review of individual patient records with a graphical user interface |
| Conduct observational studies | Support tools for observational healthcare research |
| Conduct predictive studies using machine learning | Support tools for predictive studies using machine learning |
| Add new functionalities | Capability to extend and customize the platform with new features |
| Inter-user communication module | Features for direct communication and collaboration between users |
| Integrated git module | Version control integration for code management and collaboration |

*This table lists the specifications expected for software development.*

### Development and technical implementation

The software was created by healthcare professionals (BD, AP, BP, TS) and data scientists (AL), ensuring development choices to be validated so as to meet the expectations of both stakeholders.

The software was developed with R, a programming language widely used in statistical computing and data science (13). The user interface was implemented with *Shiny*, an R package, which enables the swift creation of web applications without requiring advanced knowledge of HTML, CSS and JavaScript (14). The user interface was developed using the *shiny.fluent* library (15). Authentication within the application was managed through the *shinymanager* library (16). Editing of git repositories was managed using the *git2r* package (17).

The application was a product of the French association InterHop and was released under the GPLv3 license.

## Results

### Overall organization

LinkR operates as a collaborative platform, facilitating teamwork both locally and remotely. Locally, it enables data scientists and healthcare professionals to work together through a unified graphical user interface alongside a complete R and Python programming environment. Remotely, it facilitates the sharing of work, including study source code, data cleaning scripts, plugins, and more.

The initial step in a LinkR project begins with data importing, followed by user-initiated study creation. Each study is organized into various tabs containing widgets. Plugins, written in R using the *Shiny* library, enhance the application by adding new features, including graphical interfaces for specific tasks such as data visualization in histogram form or conducting statistical analyses. Widgets, meanwhile, allow users to display and analyze selected data in specifics ways, by applying plugins to the data. Therefore, widgets represent the execution of plugins code with the concepts selected by the user. The user repeats this widget creation action, selecting the plugin and concepts for each stage of his study or chart review.

### Data import and study creation

Data can be imported from data files such as CSV, Excel or Parquet, or from a database connection.

Following data import, users can create studies based on specific inclusion criteria and divide these into subsets based on specific patients’ characteristics, such as those with a particular medical history (*Figure 1*).

**Figure 1.**
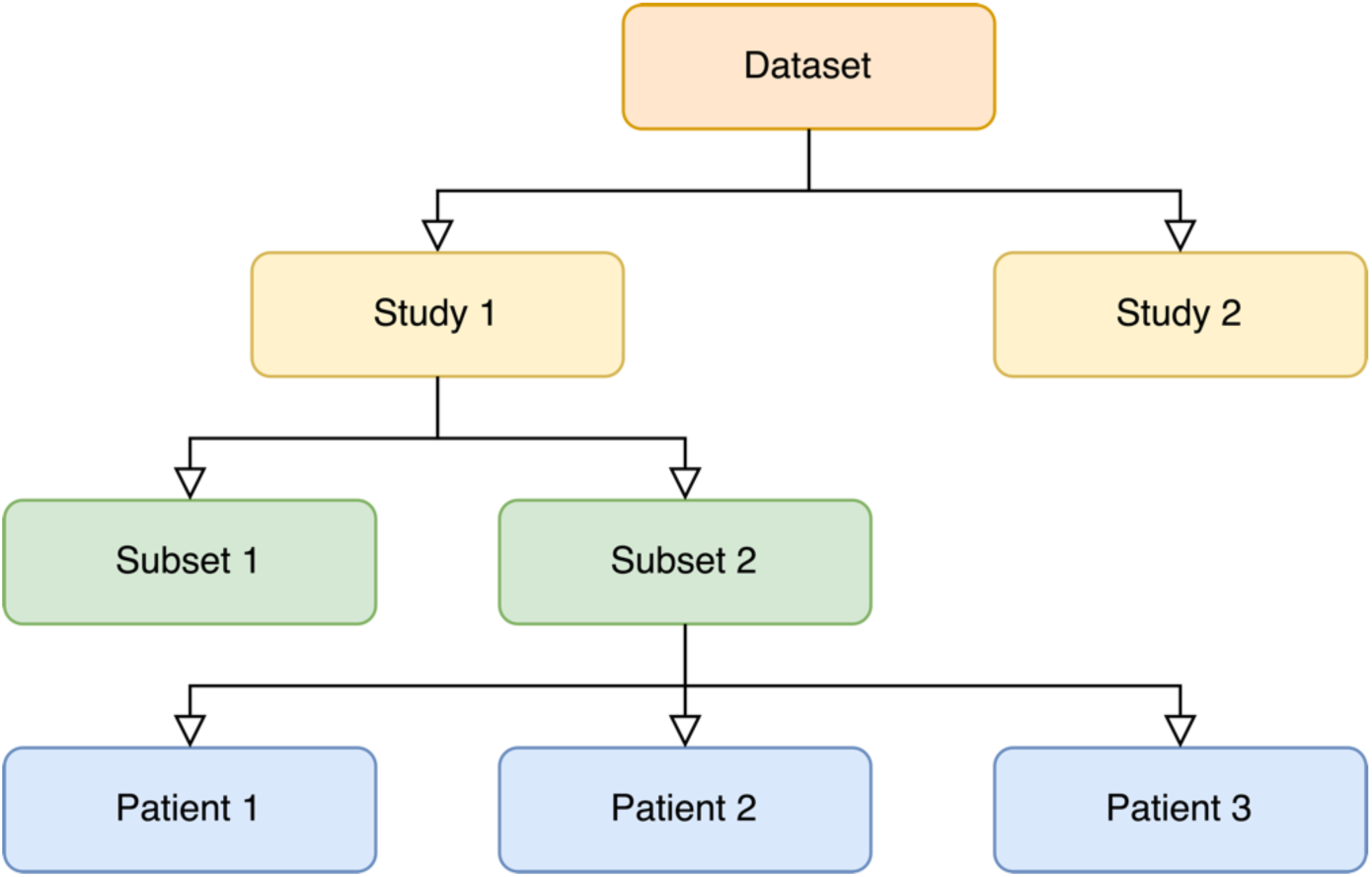
Data structure within the application. A dataset constitutes the data imported into the software from one source. This dataset can be segmented into various studies. Each study encompasses multiple subsets, each consisting of patients grouped by distinct characteristics.

### Data visualization and analysis

The platform enables data exploration at both individual (or patient) level and population (or aggregated) levels. The data pages are segmented into three parts: a side panel, a menu and a main panel (*Figure 2*). Users select datasets, studies and subsets in the side panel. For the individual data section, they then choose a patient from the selected subset and decide which of this patient’s hospital stay should be displayed. In the menu, users can add tabs, and choose which one to display. Upon selecting a tab, they can add widgets that will appear on the main panel.

**Figure 2:**
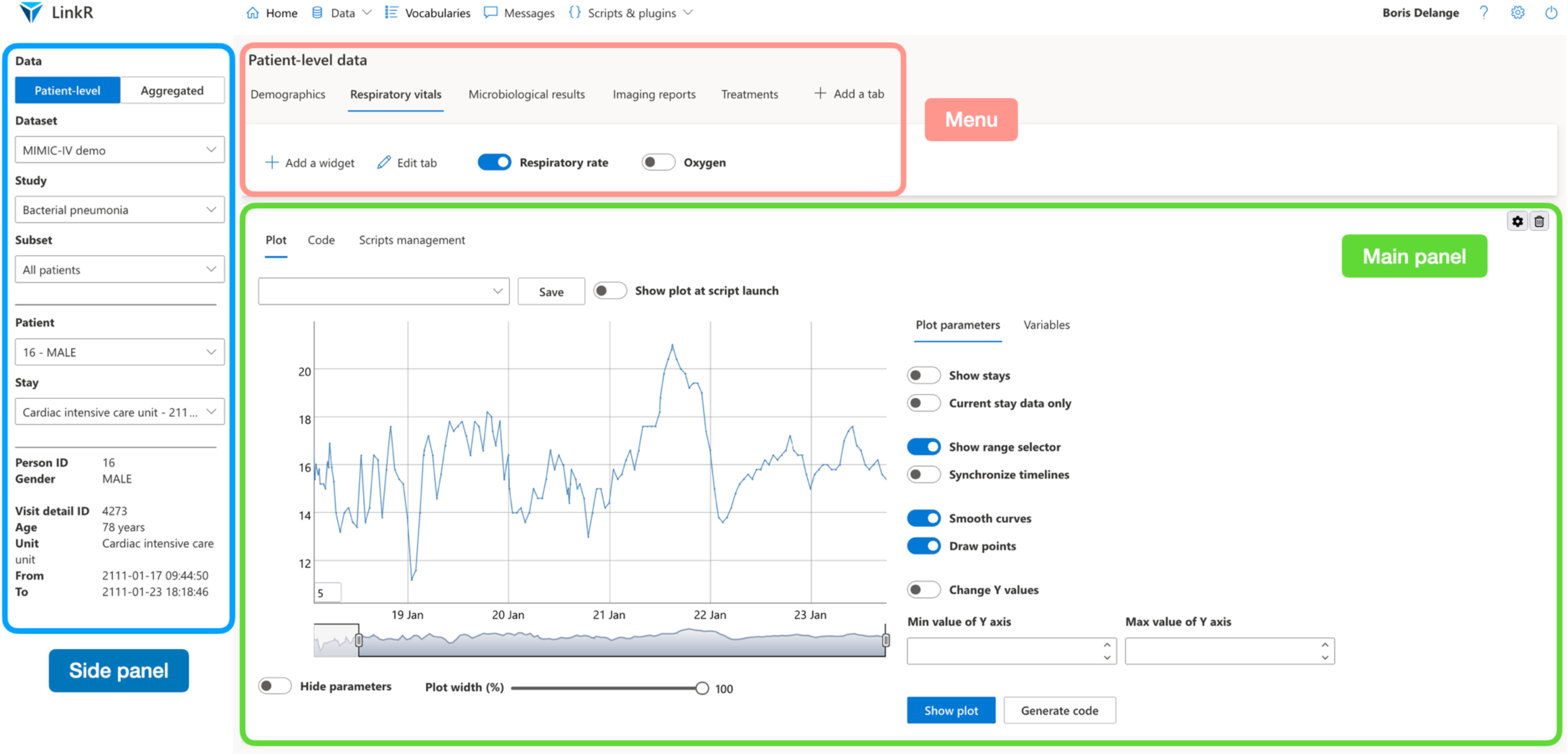
Data pages structure. *Data pages are divided into three parts: a side panel, a menu and a main panel*.

For example, *Figure 3* illustrates the use of a timeline plugin, which leverages the *dygraphs* R library to visualize data as a timeline. This plugin is applied to the ‘respiratory rate’ concept, and then displays a timeline of selected patient’s respiratory rate.

**Figure 3:**
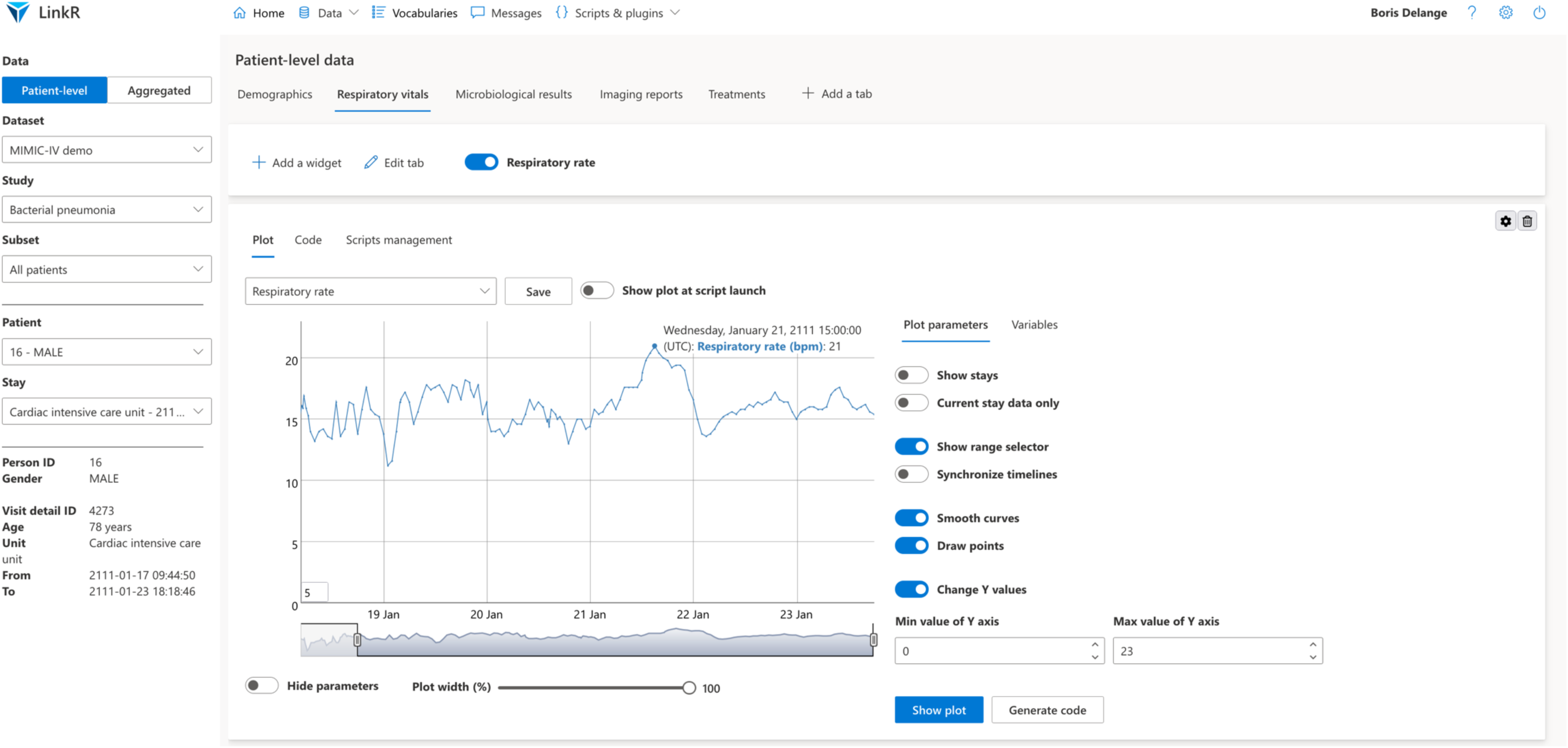
Individual (patient-level) data page. *This screenshot illustrates the use of a plugin to display the selected patient’s respiratory rate over time*.

A plugin can be reused in various contexts. For instance, a chart displaying a histogram for heart rate can be applied to another concept, such as respiratory rate (*Figure 4*).

**Figure 4:**
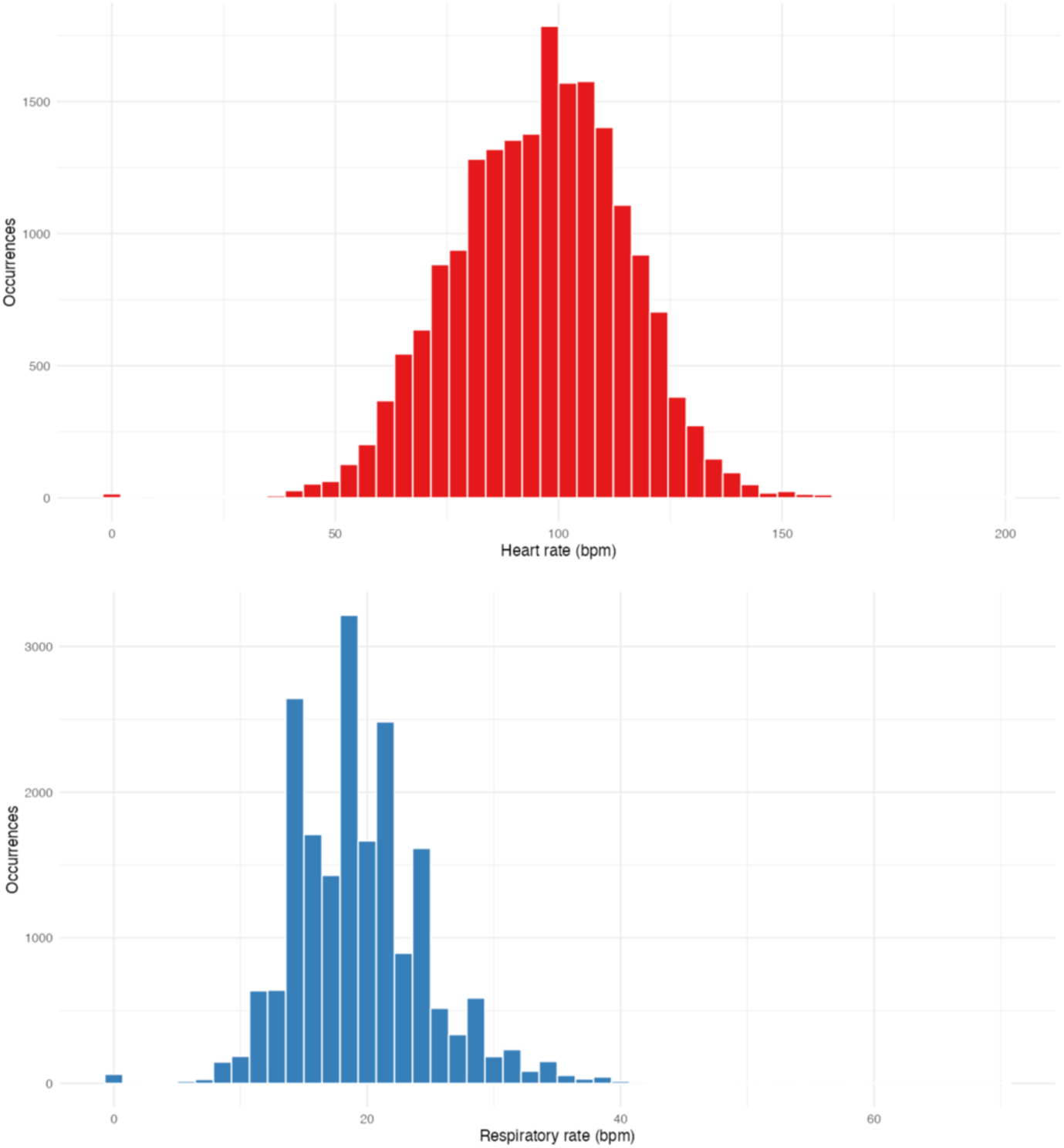
Population (aggregated) data page plugin. *This screenshot illustrates the use of a plugin to display the distribution of heart rate and respiratory rate among a group of patients*.

Plugins enable the rapid generation of code directly from the graphical user interface, which can subsequently be edited as needed (*Figure 5*).

**Figure 5:**
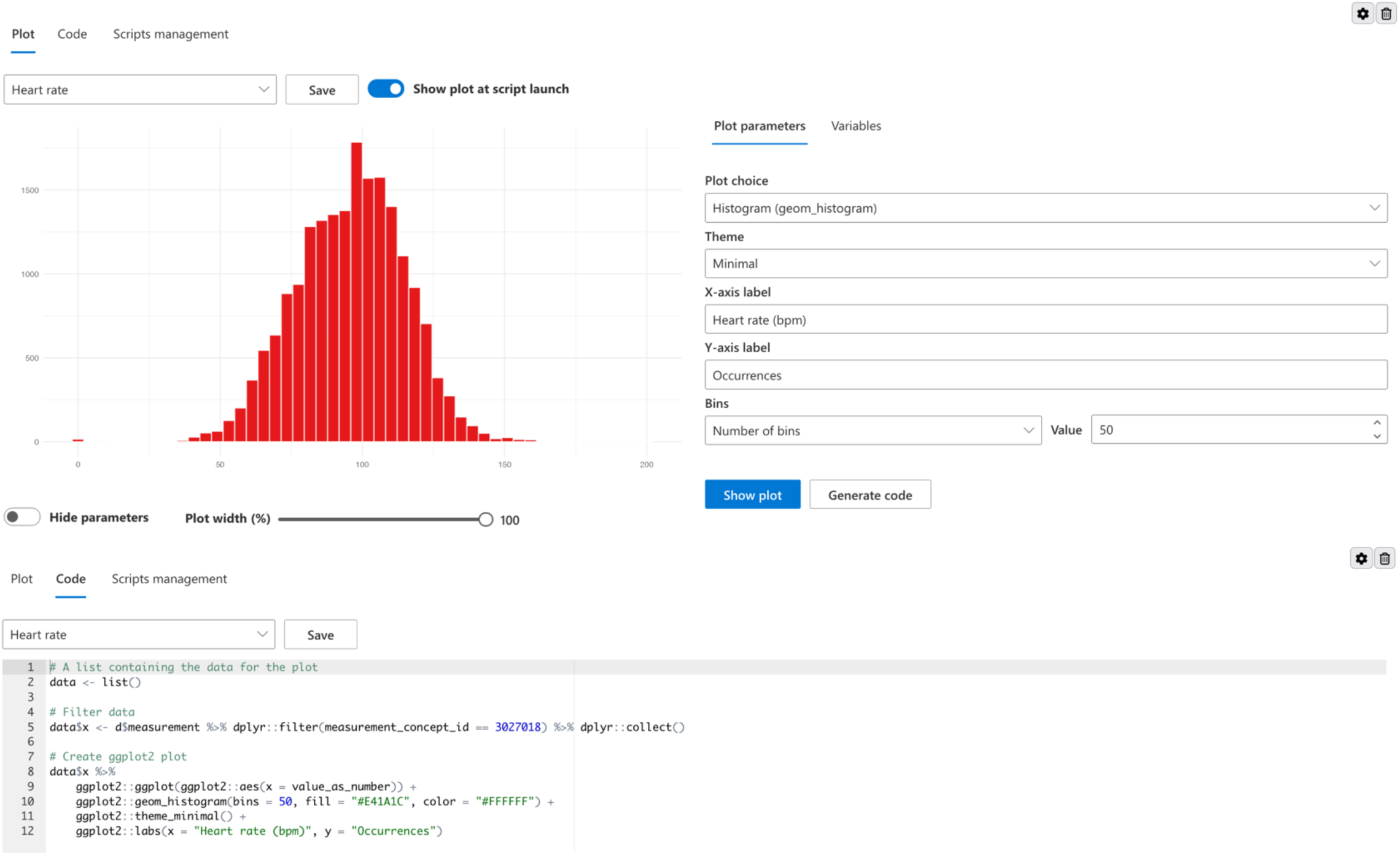
Code generation. *This screenshot highlights the plugin’s functionality in generating code that corresponds to the figure created using the graphical interface*.

On the individual data page, users create a medical record template tailored to the needs of their study. For example, if the study focuses on bacterial pneumonia, the user might create a tab displaying medical imaging reports, another for bacteriological results, and a third for the antibiotic treatments administered to the patients. When another patient is selected from the dropdown menu, the displayed data is updated accordingly (*Figure 3*).

On the population data page, users design tabs corresponding to the various stages of their study. For instance, a user can create a tab for exploratory data analysis, another for outliers exclusion, a tab to display the flowchart, and yet another to draft the study report, among others, according to their specific needs.

Thus, on one side, healthcare professionals can visualize patient data in the familiar form of a medical record, on the other side, data scientists can modify the widgets’ code if they wish, and last, students can learn how to manipulate medical data.

### Open science and collaborative work

LinkR facilitates the sharing of all work conducted within the application, such as data cleaning scripts, studies, plugins, scripts for importing datasets and terminologies, thanks to an integrated git module, using the *git2r* library. This enables the user to update gitlab or github repositories directly from within the software (*Figures 6 and 7*).

**Figure 6:**
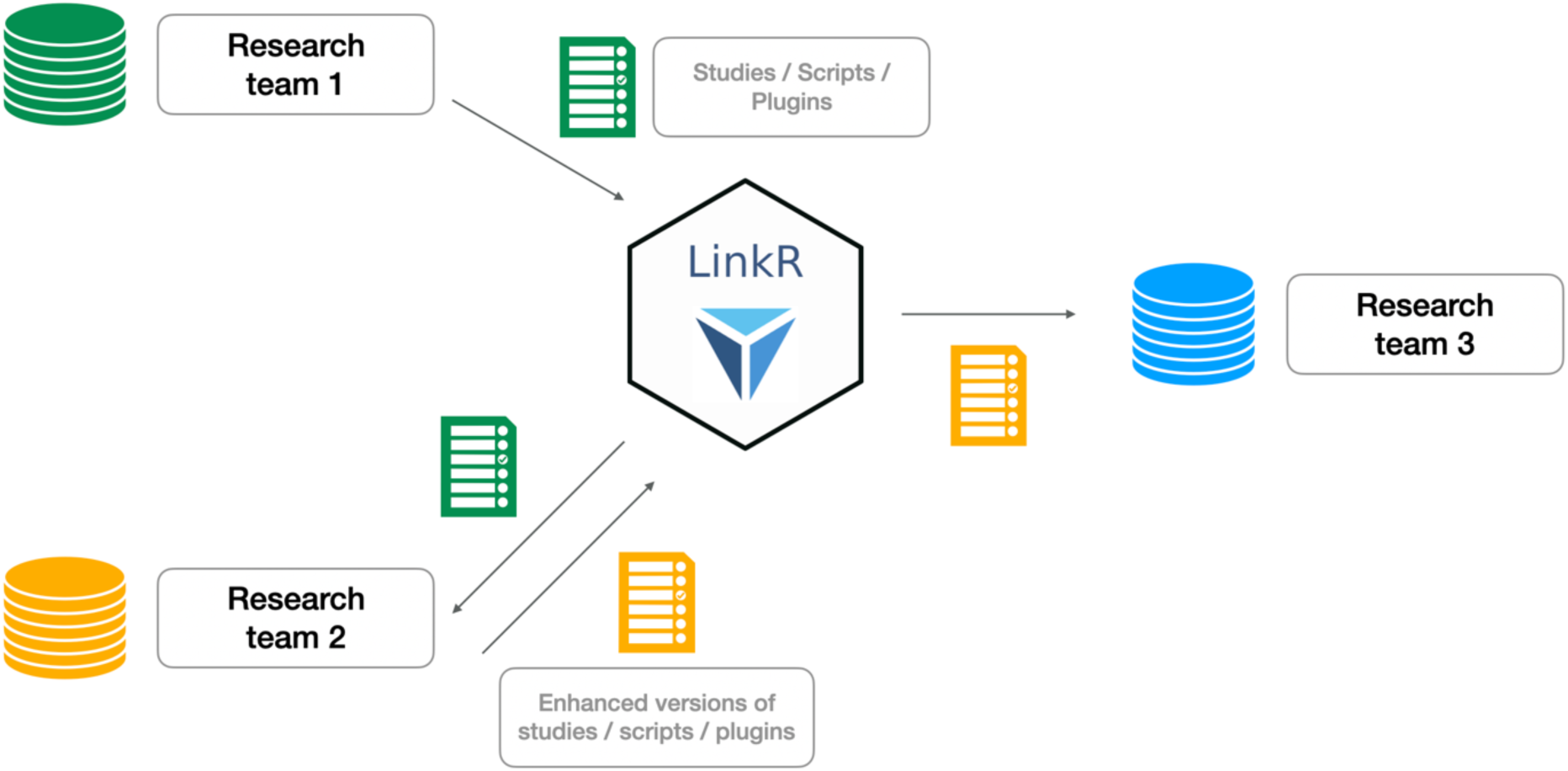
Collaborative work using LinkR. *A diagram depicting data sharing among various research teams through the software’s git module*.

**Figure 7:**
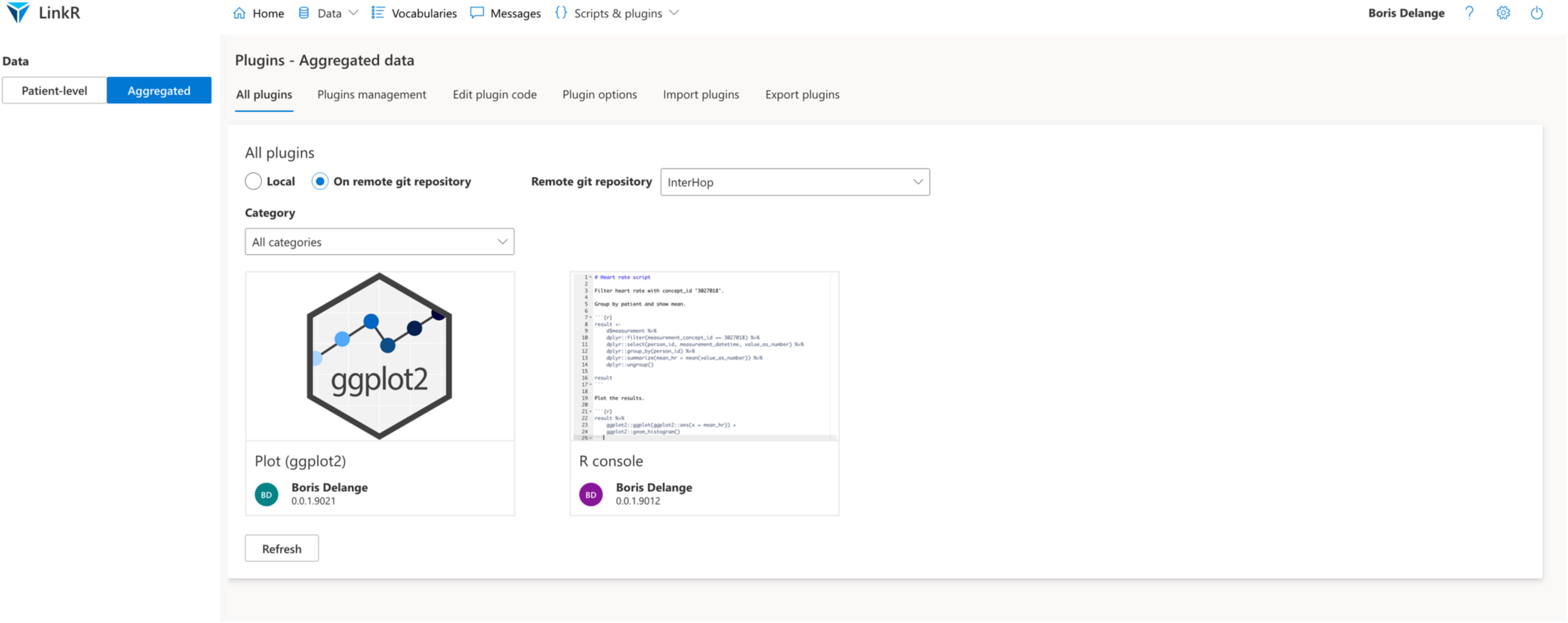
Plugins catalog. *This screenshot displays the plugins catalog where users can browse and access plugins available on another research center’s git repository. It allows users to read the descriptions of plugins and download the plugins to their local LinkR instance*.

A messaging interface facilitates collaboration among various users (*Figure 8*). For instance, a healthcare professional encountering programming challenges can share his code and seek assistance from a data scientist. Similarly, the data scientist can inquire about medical aspects from the healthcare professional. Additionally, students can ask questions to either the data scientist or the healthcare professional, depending on their specific needs. The application uses RMarkdown (18), which displays the results of R scripts within a Markdown document.

**Figure 8:**
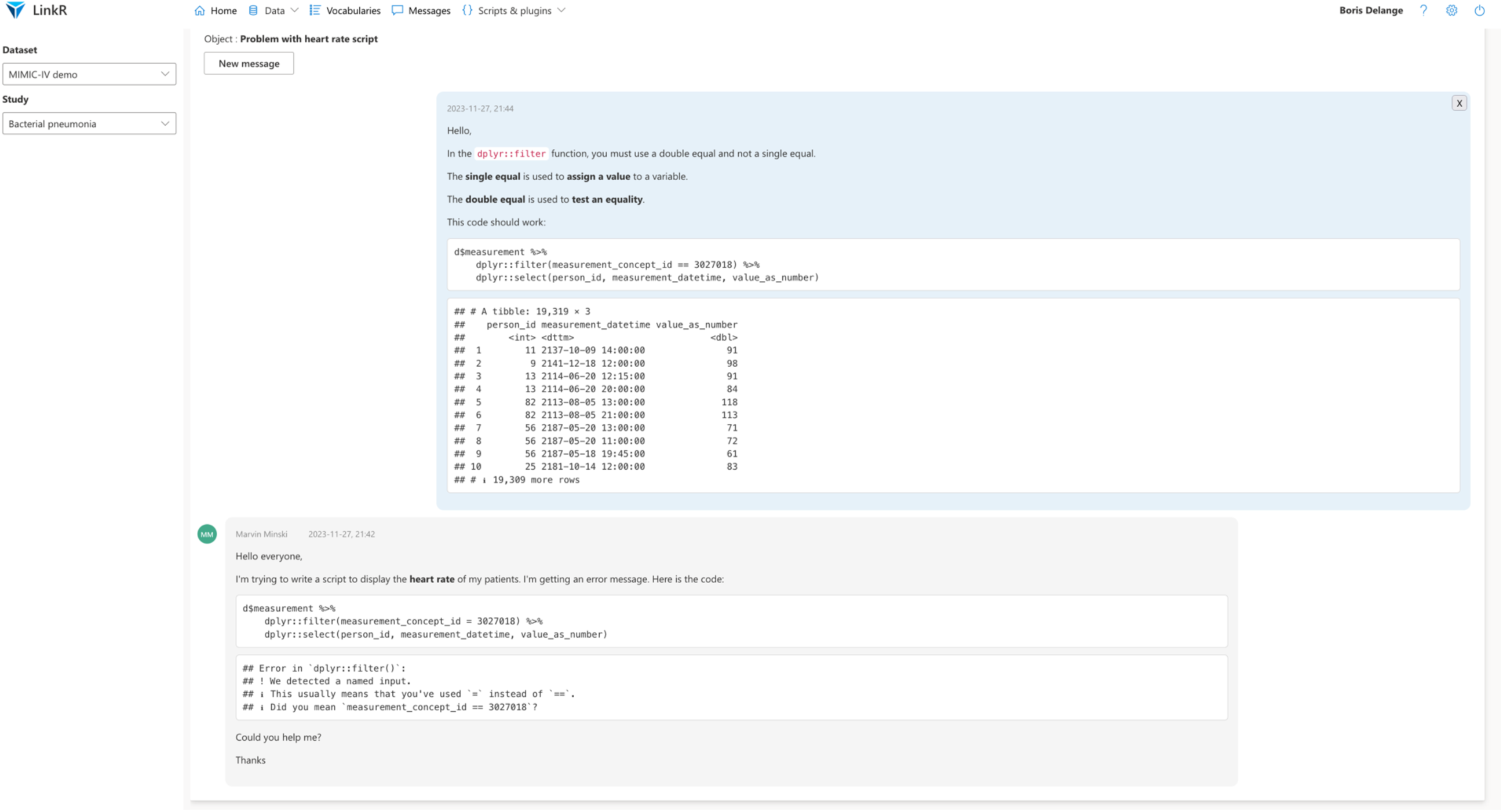
Messages page. *This screenshot displays the messages page, providing a platform for users to communicate and enhance collaborative efforts*.

### Python environment

Scripts written in Python can be executed using the *reticulate* library (19), allowing for creation of plugins using both R and Python, exploiting the full potential of both languages’ libraries.

### Help pages and tutorials

LinkR includes context-specific help pages and tutorials updated directly from the project’s Git repository. This includes data science and programming tutorials, enabling healthcare professionals and healthcare students to learn more about these topics.

### Application deployment

The application can be deployed on personal computers as well as on servers, using platforms like Shiny Server (20) or ShinyProxy (21). In the case of ShinyProxy, the application is deployed through a Docker image, simplifying the deployment process and enabling additional applications like Jupyter Notebook and RStudio to be integrated into the same production environment.

### Code availability

The code and an installation tutorial are available on this link: https://framagit.org/interhop/linkr/linkr.

## Discussion

We propose an innovative health data science platform, designed to democratize access to health data for scientists while embracing open science principles.

This low-code software empowers healthcare professionals to access data through a graphical interface that requires no programming knowledge. It also speeds up data scientists’ workflow through the sharing of data cleaning scripts and study codes. Finally, it introduces students to patient data manipulation and data science in medicine.

The sharing of data cleaning scripts enhances datasets utilization through the continual refinement of data cleaning methods across studies, leading to an ongoing improvement in the relevance of results. Moreover, sharing study codes facilitates the process of verifying the reproducibility of studies across diverse datasets.

As noticed by Lehne *et al.* (2), the current medical landscape seems less characterized by “big data” but rather by numerous disconnected small datasets, mainly due to the lack of interoperability between IT systems and clinical data warehouses. Interoperable and standardized data are essential for improving medical research efficiency and unlocking the full potential of big data in medicine. Our software specifically addresses this need for interoperability.

Other comparable tools have been developed, ATLAS being one example created by OHDSI. This is an open-source software tool for analyzing data in OMOP format, creating cohorts, performing statistical analyses, and visualizing patient medical records. However, ATLAS lacks direct R or Python programming capabilities within the software, nor does it facilitate the incorporation of additional functionalities. Moreover, it does not offer customization options for displaying medical records, which are crucial for healthcare professionals. Compared to RStudio and Jupyter Notebook, our software provides a graphical interface for data manipulation, specifically aimed at users without programming skills. *Table 2 and Figure 9* summarizes the functionalities provided by the various software packages mentioned above.

**Figure 9:**
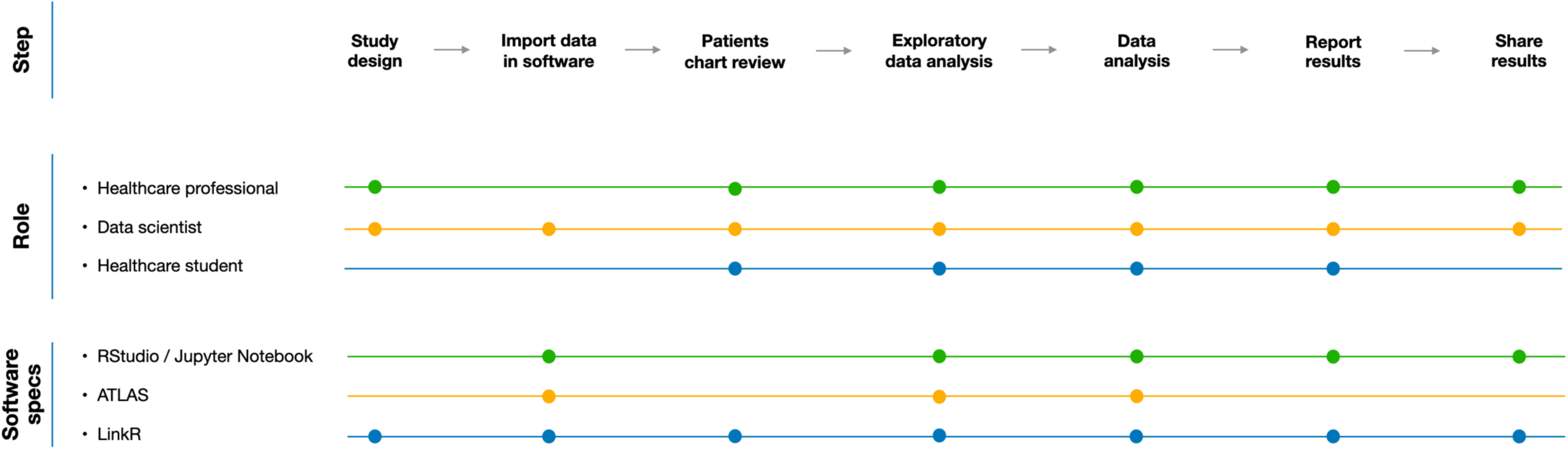
Technical specifications for different data science software packages. *This figure outlines the technical specifications of different data science software packages, according to the stages involved in carrying out a study*.

**Table 2.** Comparison of data science software and LinkR features.

| Feature | RStudio | Jupyter Notebook | ATLAS | LinkR |
| --- | --- | --- | --- | --- |
| Import data from various sources | X | X |  | X |
| Use a common data model | X | X | X | X |
| Graphical user interface |  |  | X | X |
| R programming interface | X | X |  | X |
| Python programming interface | X | X |  | X |
| Patients chart review |  |  |  | X |
| Conduct observational studies | X | X | X | X |
| Conduct predictive studies using machine learning | X | X | X | X |
| Add new functionalities | X | X |  | X |
| Inter-user communication module |  |  |  | X |
| Integrated git module | X | X |  | X |

*A cross means that the software has this feature.*

However, our study has certain limitations. Firstly, the software depends on a R backend with multiple dependencies. Secondly, in its current early stages of development, it does not allow complete studies to be carried out using a graphical interface alone and still requires advanced programming skills. Finally, the software has not been tested on a large scale.

The next step of our project is to create a web application with a Python backend, which generally performs better than R, particularly for large datasets. LinkR, currently based on the OMOP data model, will aim to integrate various common data models, including FHIR (5). The integration of large language models directly into the application could facilitate study creation and assist with R and Python programming.

LinkR, by enabling the sharing of study code, allows for the code to be executed in a decentralized manner, without the need to mobilize the data. The subsequent phase involves developing plugins for federated data analysis, using solutions such as DataSHIELD or PySyft (22,23).

The software will undergo deployment and testing with diverse user groups, each having varying degrees in programming and data science experience. User feedback will be used to refine and improve the software’s capabilities and user experience.

## Conclusion

LinkR is a low-code data science platform that democratizes access, manipulation, and analysis of data from clinical data warehouses and facilitates collaborative work on healthcare data, using an open science approach.

## Data Availability

All the source code of the application presented in this article is available as open source.

https://framagit.org/interhop/linkr/linkr

## Authors’ Contributions

BD conceptualized the study, participated in its design, developed the software’s source code, conducted the literature research and drafted the manuscript. AP, BP, AL and TS conceptualized the study, participated in its design and assisted in drafting the manuscript. All authors have read and approved the final manuscript.

## References

1. Coorevits P, Sundgren M, Klein GO, Bahr A, Claerhout B, Daniel C, et al. Electronic health records: new opportunities for clinical research. J Intern Med. 2013 Dec;274(6):547–60.

2. Lehne M, Sass J, Essenwanger A, Schepers J, Thun S. Why digital medicine depends on interoperability. Npj Digit Med. 2019 Aug 20;2(1):1–5.

3. OMOP Common Data Model [Internet]. [cited 2023 Jun 28]. Available from: https://ohdsi.github.io/CommonDataModel/

4. Stang PE, Ryan PB, Racoosin JA, Overhage JM, Hartzema AG, Reich C, et al. Advancing the science for active surveillance: rationale and design for the Observational Medical Outcomes Partnership. Ann Intern Med. 2010 Nov 2;153(9):600–6.

5. Index - FHIR v5.0.0 [Internet]. [cited 2023 Jun 28]. Available from: https://hl7.org/FHIR/index.html

6. Levin N, Leonelli S, Weckowska D, Castle D, Dupré J. How Do Scientists Define Openness? Exploring the Relationship Between Open Science Policies and Research Practice. Bull Sci Technol Soc. 2016 Jun;36(2):128–41.

7. RStudio Team. RStudio: Integrated Development for R. 2020; Available from: http://www.rstudio.com/

8. Project Jupyter [Internet]. Available from: https://jupyter.org

9. Morton CE, Smith SF, Lwin T, George M, Williams M. Computer Programming: Should Medical Students Be Learning It? JMIR Med Educ. 2019 Mar 22;5(1):e11940.

10. SAS: software and solutions for Analytics, AI & Data Management [Internet]. [cited 2024 Jan 15]. Available from: https://www.sas.com/fr_fr/home.html

11. IBM SPSS Statistics [Internet]. [cited 2024 Jan 15]. Available from: https://www.ibm.com/fr-fr/products/spss-statistics

12. Software Tools – OHDSI [Internet]. [cited 2023 Dec 21]. Available from: https://www.ohdsi.org/software-tools/

13. R Core Team. R: A Language and Environment for Statistical Computing [Internet]. Vienna, Austria: R Foundation for Statistical Computing; 2018. Available from: https://www.R-project.org/

14. Chang W, Cheng J, Allaire J, Sievert C, Schloerke B, Xie Y, et al. shiny: Web Application Framework for R. R package version 1.8.0.9000 [Internet]. [cited 2023 Nov 27]. Available from: https://shiny.posit.co/

15. Żyła K, Sobolewski J, Rogala M. shiny.fluent: Microsoft Fluent UI for Shiny Apps. 2023.

16. Thieurmel B, Perrier V. shinymanager: Authentication Management for “Shiny” Applications [Internet]. 2023. Available from: https://github.com/datastorm-open/shinymanager

17. git2r: Provides Access to Git Repositories [Internet]. 2024. Available from: https://docs.ropensci.org/git2r/ (website)

18. rmarkdown: Dynamic Documents for R [Internet]. 2023. Available from: https://github.com/rstudio/rmarkdown

19. Ushey K, Allaire JJ, Tang Y. reticulate: Interface to “Python.” 2023.

20. rstudio/shiny-server [Internet]. RStudio; 2024 [cited 2024 Jan 19]. Available from: https://github.com/rstudio/shiny-server

21. ShinyProxy [Internet]. [cited 2024 Jan 19]. Available from: https://www.shinyproxy.io/

22. Gaye A, Marcon Y, Isaeva J, LaFlamme P, Turner A, Jones EM, et al. DataSHIELD: taking the analysis to the data, not the data to the analysis. Int J Epidemiol. 2014 Dec;43(6):1929–44.

23. OpenMined - Building AI governance infrastructure to facilitate AI safety research across the globe [Internet]. [cited 2024 Jan 16]. Available from: https://openmined.org

